# Real-world performance and safety of Halova^®^ ovules in reducing the vaginal symptoms associated with vulvovaginal atrophy and postmenopausal sexual dysfunction

**DOI:** 10.1101/2023.05.17.23290093

**Authors:** Dominic-Gabriel Iliescu, Ramona Petrita, Cristina Teodorescu, Raluca Alexandra Olaru, Andreea Anda Alexa, Izabella Petre

## Abstract

Decreasing sex hormones levels during the postmenopausal period results in tissue atrophy and physiological changes, such as thinning of the vaginal epithelium, prolapse, and decreased pelvic floor strength and control. Sexual dysfunction associated with vaginal dryness has frequently been reported in postmenopausal women. The present study was designed as an observational, multicenter, real-world clinical investigation to evaluate the performance and safety of the medical device Halova^®^ ovules in reducing vaginal symptoms associated with vulvovaginal atrophy and sexual dysfunction. A total of 249 female participants were treated with Halova^®^ ovules, both in monotherapy and in combination with concomitant medication. The primary objective of this study was to evaluate the tolerability of Halova^®^ ovules in the management of symptoms associated with perimenopause or genitourinary syndrome of menopause. The evolution of clinical manifestations such as vaginal dryness, dysuria, dyspareunia, and endometrial thickness in patients with the aforementioned conditions was defined as a secondary objective. Halova^®^ ovules were rated with “excellent” clinical performance by 92.74% of the study participants as a standalone treatment and 95.71% of the study participants when used in association. Sexual dysfunction-related parameters, such as vaginal dryness and dyspareunia, were reduced by similar percentages in each arm. No adverse reactions related to treatment with Halova^®^ have been reported. Halova^®^ may be a therapeutic alternative for patients with controversial local estrogenic treatment. The study and its details are registered with NCT05654610.

## 1. Introduction

### Prevalence and etiology of vulvovaginal atrophy

Vaginal dryness can be a warning sign of vulvovaginal atrophy, also known as the genitourinary syndrome of menopause. Vulvovaginal atrophy has important medical and psychological consequences and is the most common problem during menopause. Unlike hot flashes and night sweats, which resolve spontaneously, the symptoms of vaginal dryness affecting the lower urinary tract develop over time and are very distressing. The prevalence of vaginal dryness increases in the years following menopause and causes bothersome symptoms such as itching, burning, and pain during intercourse. Furthermore, it is estimated that approximately 17% of women between the ages of 17 and 50 have difficulties with successful sexual contact due to vaginal dryness and pain during sexual contact, which leads to anxiety and decreased libido over time (1). Its prevalence ranges from 36% to almost 84%, and it is often underdiagnosed and undertreated (2,3). The condition may also occur earlier, in perimenopausal women who take antiestrogenic medications or who have medical characterized by decreased levels of estrogen (4).

Currently, estrogen therapy, which is approved for the treatment of vaginal atrophy, is often associated with adverse effects and multiple contraindications in menopausal women. Among these, metabolic imbalances, mood swings, bloating, and risk of developing ovarian cancer are the most common. (5,6) Furthermore, long-term use of estrogen therapy may lead to breast cancer and should therefore be limited. (7) During the reproductive age, ovaries produce a large proportion of circulating estrogens (4,7). Normally, the vaginal walls are lubricated by cervical-vaginal fluid produced by the cervix at the top of the vagina. Decreasing sex hormone levels during the postmenopausal period reduces sexual desire and affects all layers of the vagina, leading to narrowing of the vagina, loss of rugae, keratinization of the surface, and thinning of the vaginal epithelium. A thin vaginal epithelium leads to increased susceptibility to trauma, with clinical signs such as bleeding, petechiae, and ulceration with any type of pressure, including sexual activity or a simple gynecological maneuver. (4,8).

In addition to the onset of menopause, estrogen levels can decrease significantly due to other causes such as birth or breastfeeding, cancer treatment, surgical removal of the ovaries, and anti-estrogen drugs used to treat uterine fibroids and endometriosis. Other causes of vulvovaginal atrophy include Sjogren’s syndrome, allergies, cold medicines, antidepressants, vaginal washings, anxiety, and stress overload (7,9).

### Clinical manifestations

The main symptoms of vulvovaginal atrophy include vaginal dryness and reduced lubrication during sexual activity, urge incontinence inflammation, itching, discomfort, atypical vaginal discharge, and dyspareunia, which greatly affect quality of life (10). In addition, recurrent tract infections have frequently been reported (6).

### Clinical management of urinary complaints

Urinary complaints in postmenopausal women should be managed following physical examination and laboratory diagnostic testing, including serum hormone levels and Papanicolaou smear. Differential diagnosis should be considered to eliminate any vaginal infection (candidiasis, trichomoniasis, or bacterial vaginosis) or other conditions that cause chronic vaginal and vulvar itching, discharge, or pain such as irritants and vulvovaginal dermatoses (11). Irritants that can cause chronic vaginal itching include perfumes, locally applied lubricants or cosmetic soaps. Vulvovaginal dermatoses that may cause similar symptoms include lichen sclerosus, lichen planus, and lichen simplex chronicus.

The principal therapeutic target in the management of vaginal atrophy is the relief of symptoms, particularly vaginal dryness. Treatment strategies are mainly based on the use of moisturizers and lubricants, including physical therapy, low-dose vaginal estrogen therapy, vaginal dehydroepiandrosterone, and oral ospemifene, with a more modern approach including the use of vaginal lasers (12,13). Patients who experience problems with natural vaginal lubrication due to hormonal changes often benefit from estrogen therapy. In addition to estrogen treatment, or if side effects caused by hormone treatment are difficult to bear, preparations with moisturizing agents that help introduce and maintain water in the vaginal mucosa are recommended.

Non-hormonal treatments include vaginal/topical moisturizers and lubricants, such as a combination of Hippophaë rhamnoides oil, hyaluronic acid, and glycogen or hyaluronic acid, collagen, isoflavones, and vitamins (14,15). Lubricants provide short-term relief for patients and are typically used for vaginal dryness during intercourse, whereas moisturizers have long-lasting effects and may be used daily every 2-3 days per week (16).

Hormonal replacement therapy may be systemic (oral estrogen replacement) or localized (intravaginal/topical estrogen, intravaginal releasing rings, and vaginal dehydroepiandrosterone). Estrogenic therapy is considered the most effective treatment for vaginal atrophy, dryness, and dyspareunia in women with estrogen deficiency. On the other hand, estrogen therapy is known for its increased risk of stroke and thromboembolism. In addition, estrogen therapy should be administered with caution in patients who survive hormone-sensitive cancers, as the systemic absorption of estrogen can stimulate the growth of breast cancer cells. Although systemic estrogen therapy improves symptoms of vaginal atrophy, systemic doses are higher than those used for topical application and should be administered only if other menopausal symptoms requiring treatment are present (4,8,17).

Post-marketing studies are primarily used to answer the important clinical question, “Is this medical device effective and safe in a non-controlled, real-life setting? The primary objective of this study was to observe the tolerability of Halova^®^ ovules in treating vaginal dryness and restoring the natural lubrication of the vaginal mucosa. The secondary objective of this study was to evaluate the performance of the medical device by clinical examination in reducing the symptomatology correlated with vulvovaginal atrophy, evaluation of endometrium thickness, and vaginal pH. Additionally, the degree of patient satisfaction related to the use of a medical device (Likert scale) was assessed.

## 2. Methods

### Study design

The present study was designed as part of a medical device post-marketing clinical follow-up, involving routine care from a variety of clinical practices. The study had an open-label, multicenter, non-randomized, real-world evidence study design. The data were collected between the 1^st^ of March 2022 and the 31^st^ of July 2022. The clinical sites and locations are listed in table 1.

**Table 1.**
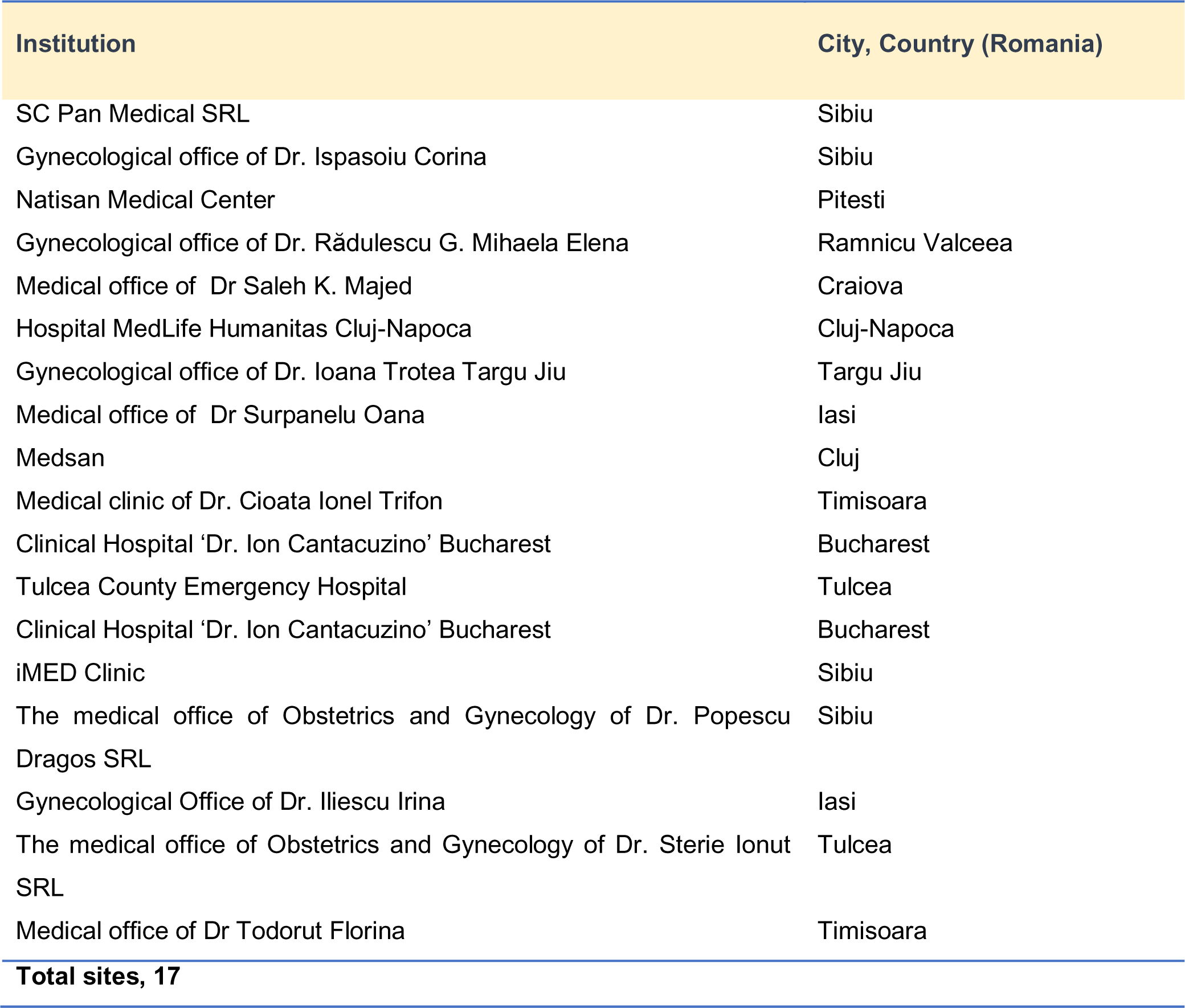
Clinical practices and their locations.

### Participant population

The participant population included women aged between 18-70 years, of Caucasian ethnicity with several clinical manifestations associated with the following conditions: dryness in the vaginal region, perimenopause, vulvovaginal atrophy, menopausal disorder, or vaginal prolapse. Two hundred forty-nine women were evaluated. One hundred seventy-nine patients had only undergone treatment with Halova^®^ ovules as monotherapy, while 70 had used Halova^®^ ovules in association with other medications (polytherapy). Subjects with psoriasis, vitiligo, plantar ulcers, lipoid necrobiosis, granuloma annulare, vulvar, or cervical cancer were excluded. Based on other similar pivotal studies, the sample size initially included a total of 240 subjects, with baseline characteristics shown in table 2.

**Table 2.** Baseline demographic data for the sample population.

| Baseline characteristics | Subjects |  |
| --- | --- | --- |
|  | n | % |
| Age (years) |  |  |
| < 50 | 77 | 32.08 |
| ≥ 50 | 163 | 67.91 |
| Gender |  |  |
| Female | 240 | 100.00 |
| Ethnicity |  |  |
| Caucasian | 240 | 100.00 |
| Menopausal status |  |  |
| Premenopausal | 42 | 17.92 |
| Perimenopausal | 35 | 14.58 |
| Menopausal | 162 | 67.50 |
| Physical activity |  |  |
| Yes | 49 | 20.42 |
| No | 191 | 79.58 |
| Sexual activity |  |  |
| Yes | 141 | 58.75 |
| No | 99 | 41.25 |

The study involved 17 Romanian specialist physicians as investigators, each with 4 - 20 patients under treatment with Halova^®^ ovules. Endometrial evaluation was performed by transvaginal ultrasonography. The total duration of the study was 30 days. The medical device was applied once daily for ten consecutive days.

Prospective data were collected from each patient, including the initial diagnosis, transvaginal ultrasonography, vaginal pH value, vaginal symptoms, worsening symptoms, and adverse events. To assess pH, a piece of litmus paper was placed on the lateral vaginal wall until moistening. A pH ≥ 4.6 or greater indicated vulvovaginal atrophy, assuming the patient did not have bacterial vaginosis (if tests and wet mount were performed to exclude infection with *Gardnerella vaginalis).* Primary and secondary outcomes were collected at baseline and after 30 days.

### The medical device

Halova^®^ is a medical device manufactured by Perfect Care Manufacturing S.R.L and identified under the European Medical Device Regulation with device identification number 5944754000754. Intravaginal administration is intended to promote and accelerate hydration, healing, epithelialization, and/or soothing of the injured, atrophic, or irritated vaginal mucosa. Halova^®^ vaginal ovules are composed of sodium hyaluronate (5 mg), marigold extract (60 mg), vitamin E (10 mg), Aloe Vera oil (60 mg), semi-synthetic glycerides (1587 mg), lanolin (50 mg), silicon dioxide (25 mg), and xylitol (3 mg).

The ovules melt evenly in the vaginal mucosa, forming a cream that contributes to restoring the normal lubrication of the vaginal mucosa and helps preserve normal pH and vaginal flora. Halova^®^ ovules are intended for use in adult women (including those in menopause) and are indicated for vaginal dryness caused by age, various pathologies, or other drug treatments, relief of pain and discomfort during sexual intercourse, balancing of vaginal flora, and vaginal pH preservation. Halova^®^ ovules contain ingredients that provide beneficial properties for restoring normal lubrication of the vaginal mucosa and help preserve normal vaginal pH and flora.

Hyaluronic acid is an alternative to non-hormonal treatments for signs of vaginal atrophy and dyspareunia. Sodium hyaluronate retains a large amount of water, provides moisture to the vaginal tissue, and is an effective treatment for vulvovaginal discomfort. Hyaluronic acid is contained in the medical device Halova^®^ at a concentration of 5 mg/ovule and is used to promote hydration of the dry vaginal mucosa and re-epithelialization of damaged tissue. The mechanism of action of hyaluronic acid with a high molecular mass at the level of the vaginal mucosa is realized by the formation of an extracellular matrix with water trapped in the structure.

In the composition of the medical device, the extract of Calendula officinalis is in a quantity of 3,3%, which is used to prevent contamination with exogenous bacteria during handling of the medical device, and to prevent microbiological contamination of the fat base with gram-negative bacteria and fungi.

Xylitol is present in the medical device Halova^®^ at a concentration of 3 mg/ovule and acts as a nutritional substrate. The ovule base was composed of a mixture of fatty base, lanolin, and oily extract of Aloe Vera. Vitamin E prevents the oxidative degradation of the fatty base.

The medical device is in the form of ovules of 1.8 g each, ovoid in shape, white or pale yellow, with a smooth appearance, without spots of color or areas with agglomerated powders. In the longitudinal section, the ovules have a homogeneous appearance, without agglomeration of particles and air bubbles. The medical device was administered for 10 days to patients meeting the eligibility criteria.

### Ethical and regulatory aspects of the Study

Written consent for participation in the study was obtained from all patients. Owing to legal considerations (General Data Protection Regulation Directive effective from May 21, 2018, in all European Union countries), patients or their legal representatives have an absolute right to request that their data be removed from the study database. The IRB of ENTE CERTIFICATIONE MACCHINE reviewed the post-marketing clinical follow-up plan, including ethical considerations, and approved the data collection with approval number 1MD.

The study was conducted following the Guide to medical devices: ‘Post-market clinical follow-up studies (https://www.imdrf.org/sites/default/files/docs/imdrf/final/technical/imdrf-tech-210325-wng65.pdf) and the International Society for Pharmacoepidemiology (ISPE; 2015) Guidelines for ‘Good pharmacoepidemiology practices (GPP)’ (https://www.pharmacoepi.org/resources/policies/guidelines-08027/)

The data collection and study procedures were conducted in accordance with the ethical principles of the Declaration of Helsinki. Data were stored according to Annex E of ISO 14155:2020 (https://www.iso.org/standard/). The study and its details are registered at www.clinicaltrials.gov under ID NCT05654610.

### Statistical analysis

All statistical analyses were performed using Microsoft Excel Analysis ToolPak version 16.69.1. P<0.05 was considered to indicate a statistically significant difference. The quality and completeness of the collected data were preliminarily assessed and compared with data analysis. No study participants were involved in any violation of inclusion/exclusion criteria. To examine treatment significance over time, Fisher’s exact test was performed for categorical variables, and the Mann-Whitney U test was used to perform a comparative analysis for non-normally distributed variables.

## 3. Results

### Primary objectives

The primary objective was to evaluate the tolerability of Halova^®^ ovules in treating vaginal dryness and restoring natural lubrication of the vaginal mucosa. There have been no reported adverse reactions related to treatment with Halova^®^ after 30 days of administration, indicating a high safety profile. Some limitations of this study include the absence of a control group, short treatment follow-up period, and heterogeneity of the sample population. A CONSORT diagram is shown in Supplemental figure 1.

### Secondary objectives

The secondary objective of this clinical investigation was to evaluate the performance of the medical device by clinical examination, together with the degree of patient satisfaction related to the use of the medical device (5-points Likert scale).

To calculate the statistical significance related to the performance/improvement of the secondary objective indicators, we used a one-tailed z-test (0.05 significance level) with the first screening visit as the baseline (a null hypothesis that there is no statistically significant difference between baseline visit and the follow-up visit planned at 30 days interval). The p-values are presented in table 3.

**Table 3.** P-values for significance at 30 days.

| Type of treatment | Clinical performance | Vaginal pH | Endometrial thickness | Dyspareunia | Dysuria | Vaginal dryness |
| --- | --- | --- | --- | --- | --- | --- |
| Monotherapy | p < 0.0001 | p < 0.001 | p < 0.0001 | p < 0.0001 | p < 0.05 | p < 0.0001 |
| Polytherapy | p < 0.01 | p < 0.01 | p < 0.001 | p < 0.05 | p < 0.05 | p < 0.0001 |

### Clinical performance of the medical device

As shown in figure 2, regarding the clinical performance we observed in the monotherapy group, the treatment was rated for the majority of patients as “excellent” (84.35%) and with similar percentages, in the polytherapy group, the treatment was rated as “excellent” or “highly effective” for the majority of patients (95.71%).

### Vaginal pH level evaluation

The vaginal pH levels of the patients were evaluated after 30 days, as shown in figure 3. Normal vaginal pH was noted in 75% of patients treated with Halova^®^ ovules and in 84% of patients receiving the treatment comprising of Halova^®^ ovules and other specific medications.

### Dyspareunia symptoms

Regarding dyspareunia symptoms, patients were evaluated, during a brief interview, using a 5-point Scale, from “Absent” to “Severe” as shown in figure 3. We observed that in patients using Halova^®^ ovules as monotherapy, a significant proportion of 72 % was reported with the score “Absent”, while in patients receiving associated treatment, 48% of patients reported dyspareunia as “Absent” and 52% reported “Mild dyspareunia”.

### Endometrium thickness evaluation

Regarding endometrial thickness, ultrasound was performed on the study participants treated with Halova^®^. The results are shown in figure 3. We observed a significant proportion of patients with an endometrial thickness rating as “Absent”. “Mild” and “Moderate” endometrial thickness were encountered for 20%, respectively 4% of the study participants in the monotherapy group. In polytherapy, the percentages were similar, with 78% of patients with a rating of endometrial thickness as “Absent”, 20% with a rating of endometrial thickness as “Mild” and 2% with a rating of endometrial thickness as “Moderate”.

### Dysuria symptoms

As shown in figure 4, regarding the presence of dysuria, the highest proportion (64%) of patients reported a score of “Absent” on monotherapy. When Halova^®^ was administered as polytherapy, 78% of the study participants reported the symptom as “Absent”.

### Vaginal dryness symptoms

The results of the evaluation of vaginal dryness are presented in figure 5. The efficacy of the medical device in reducing vaginal dryness was observed in 82% and 80% of the patients, respectively.

### Degree of satisfaction after using Halova® ovules

In terms of the degree of satisfaction offered, participants in the largest population (94.38% of patients treated with Halova^®^ monotherapy rated the treatment as “Excellent”. Also 85,71% of patients treated with Halova^®^ ovules as polytherapy) rated the device as “Excellent” or “Highly effective”, 14.29% (as shown in figure 6).

## 4. Discussion

Vulvovaginal atrophy and its main symptoms (vaginal dryness and dyspareunia) are closely related to all domains of female sexuality. Vaginal dryness can be severe and distressing enough to affect daily activities, sexual desire, and normal sexual intercourse (18). Approximately 40% of the women with vaginal atrophy report dyspareunia. Dyspareunia is defined as the recurrent pain that occurs during sexual intercourse. This has negative effects on sexuality and sexual function. During menopause, women experience a plethora of symptoms that correlate with physiological changes. Both healthcare professionals and patients often find it difficult to approach the subject of sexual problems associated with menopause and vulvovaginal atrophy (19). With the administration of Halova^®^ ovules, the symptoms of dyspareunia were greatly reduced, thus favoring normal sexual function in affected patients.

The beneficial effects of reducing vaginal dryness can be explained by the local action of sodium hyaluronate. Multiple studies have confirmed the advantages of using topical sodium hyaluronate in vaginal dryness since it is a modern, safe, and well-tolerated product (20,21). Owing to its highly anionic properties, sodium hyaluronate can attract water to swell, create volume, and provide structural support, thereby acting as a topical lubricant. Sodium hyaluronate is the salt form of hyaluronic acid, with a smaller molecular structure that increases stability and resistance to oxidation. Water solubility is associated with better skin and mucosal penetration, leading to better hydration. The mechanical protection of the vaginal endothelium is ensured by its high viscosity. Sodium hyaluronate has been successfully used since 1980 for the treatment of various other diseases, such as dry eye, joint diseases, cystitis, atopic dermatitis, cataract extraction, osteoarthritis, and as a filler for skin wrinkles (22,23). It provides a high safety profile that has been previously studied in postmenopausal women (24).

The diminishing estrogenic activity, along with other factors, such as a rise in vaginal pH and alteration of the vaginal microflora, led to an increased susceptibility to urinary tract infections and vaginitis in postmenopausal women, but also in sexually active women. Therefore, postmenopausal women are at an increased risk of recurrent urinary tract infections, dyspareunia, vaginal irritation, pruritus, pain, and symptoms of urgency, frequency, dysuria, and urinary incontinence (25). Poor sleep, cardiometabolic symptoms, muscle and joint pain, and mood changes affect approximately 80% of women during diminishing estrogenic activity. (26)

A very interesting aspect of our study is that the Halova^®^ medical device is efficient in maintaining a healthy vaginal pH. The normal vaginal pH level (3.8 – 4.5) is important for its protective role in blocking yeast and bacterial multiplication. Thus, the supportive role of the medical device in preventing vaginal infections was proven based on its effective role in the correction of unbalanced vaginal pH, with 88.79% of patients in the single therapy arm and 95.35% in the add-on arm reporting normal vaginal pH.

The claimed therapeutic indications of the device are linked to the treatment of both postmenopausal and nonmenopausal vaginal dryness, relief of pain and discomfort during sexual intercourse, balance of the vaginal flora, treatment of vaginal pH disturbances, and boosting vaginal lubrication. Vitamin E, due to its rich composition in phytoestrogens, is a key element in stabilizing estrogen levels and can greatly improve menopausal symptoms including hot flashes, irritability, insomnia, dizziness, palpitations, shortness of breath, and vaginal dryness. When applied locally, it favors healing and re-epithelialization processes through its nourishing and moisturizing effects. A recent review conducted by Porterfield et al. elaborated on the evidence for vaginal vitamin E efficacy in reducing patient-reported genitourinary symptoms in healthy postmenopausal women compared with placebo or vaginal control therapy (27). Another review by Feduniw concluded that vitamin E might be an option for standard hormone therapy and may be an option to treat symptomatic women with contraindications to estrogen (28).

Particular concerns and controversies are associated with the treatment of vaginal atrophy and its symptoms in patients with breast cancer. The increasing use of aromatase inhibitors over the past few years has led to an increased incidence of vaginal atrophy, with a high impact on the quality of life of patients with breast cancer (29). However, systemic or topical hormonal therapy is contraindicated in patients with breast cancer. Thus, Halova^®^ might be an excellent therapeutic alternative in these patients and in patients in whom local estrogenic treatment remains controversial, such as in uterine cancer, patients with a history of deep vein thrombosis or pulmonary embolism, a history of stroke or myocardial infarction, or a history of blood clotting disorders (30).

## 5. Conclusions

Halova^®^ significantly alleviated symptoms such as vaginal dryness, dyspareunia, and dysuria while restoring the normal pH, thus harnessing the protective features of the vaginal microbiome. Once a day for a sequence of 10 consecutive days, Halova® may be a suitable treatment option for vulvovaginal atrophy and urogenital complaints in women of reproductive age and postmenopausal status. The medical device has proven safe and effective in alleviating most of the bothersome signs and symptoms of atrophic vaginitis in postmenopausal women, such as dysuria, dyspareunia, and vaginal dryness. Halova® may be an excellent therapeutic alternative for patients for whom local estrogenic treatment remains controversial.

The medical device has demonstrated anti-atrophic activity in the genitourinary tract, resulting in significantly improved symptoms associated with normal sexual functioning. Future research is needed to confirm its tolerability and performance in extending timeframes as well as during pregnancy. Some limitations of this study include the absence of a control group, short treatment follow-up, and heterogeneity of the sample population. This medical device also offers a promising treatment for conditions like endometriosis, where symptoms like dyspareunia, dysuria, and pelvic pain could be alleviated by the administration of Halova. Future research is needed to establish new efficacy endpoints in other conditions, e.g., endometriosis.

## Data Availability

All data produced in the present study are available upon reasonable request to the authors.

## Acknowledgments

The authors thank the study participants, who voluntarily enrolled in the study. All authors have approved the final version of the manuscript for publication. The authors also thank Alexandru-Remus Pinta for his work provided during the statistical analysis and Adrian Pocola for technical support provided during data management collection. All authors approved the final version of the manuscript to be published.

## Data sharing

Perfect Care Distribution provides access to all individual participant data collected during the trial, after anonymization. Access is provided after a proposal has been approved by an independent review committee identified for this purpose and after the receipt of a signed data-sharing agreement. Data and documents, including the clinical investigation plan, clinical study report, and blank or annotated case report forms, will be provided in a secure data sharing environment.

---

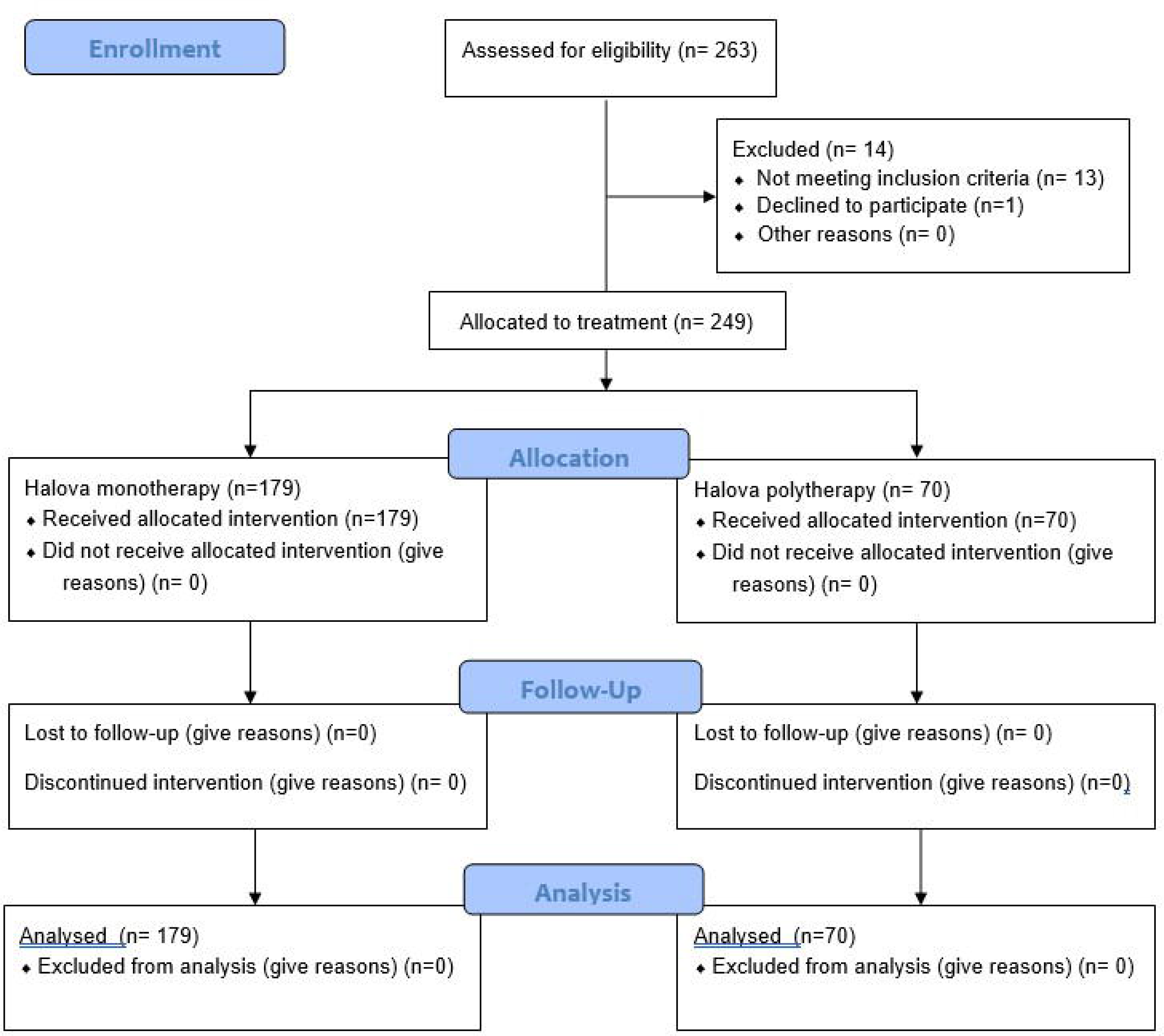

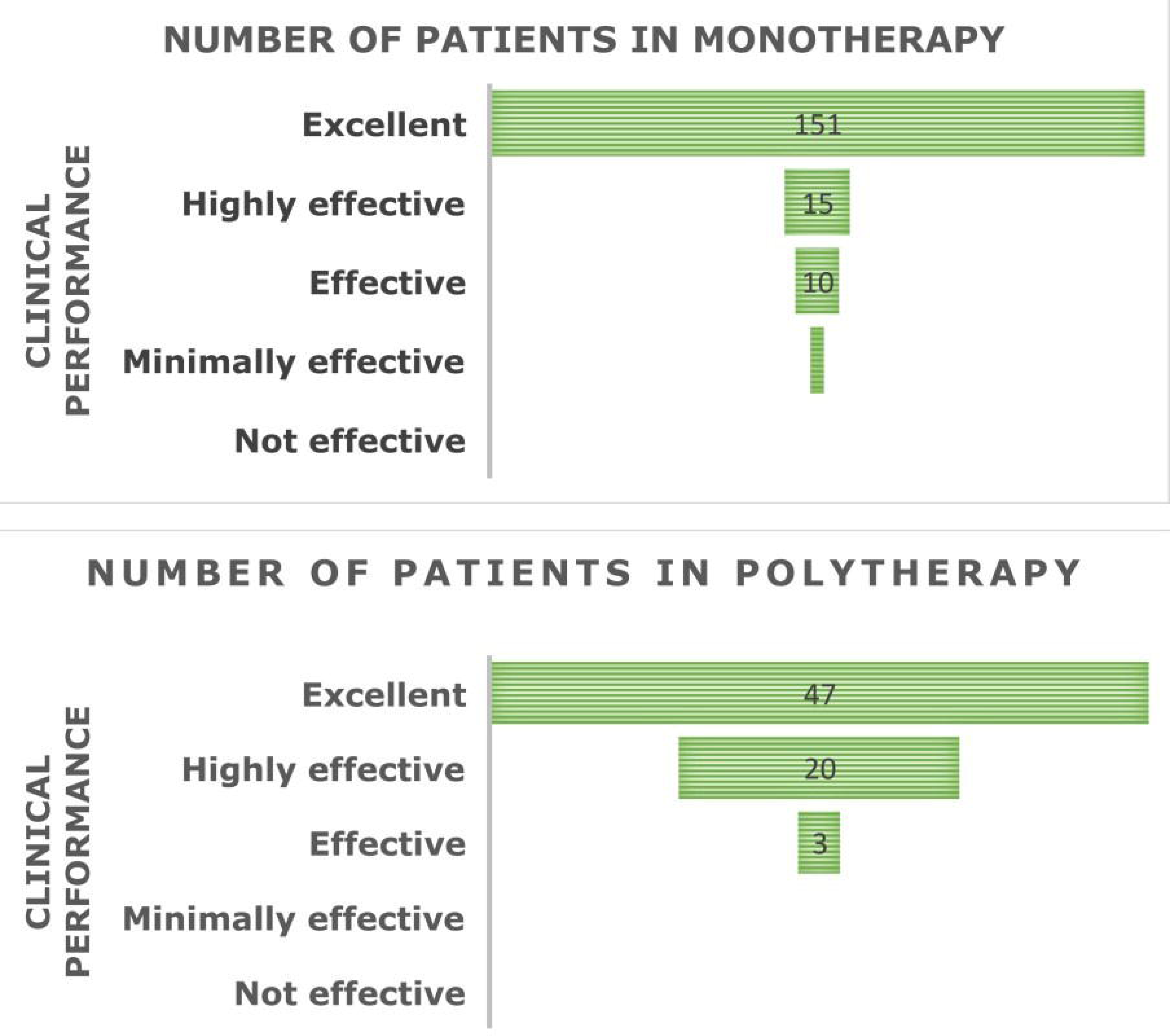

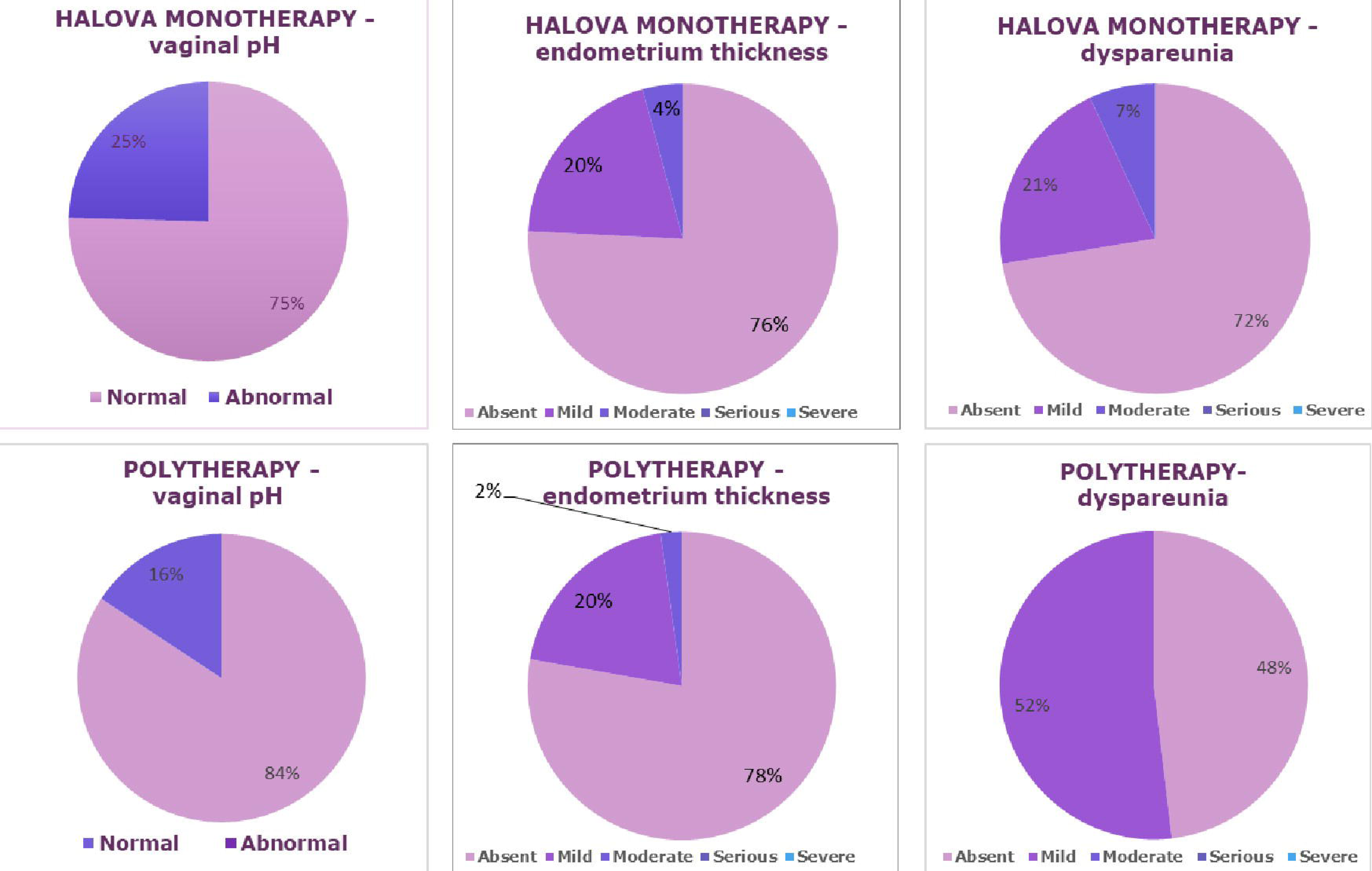

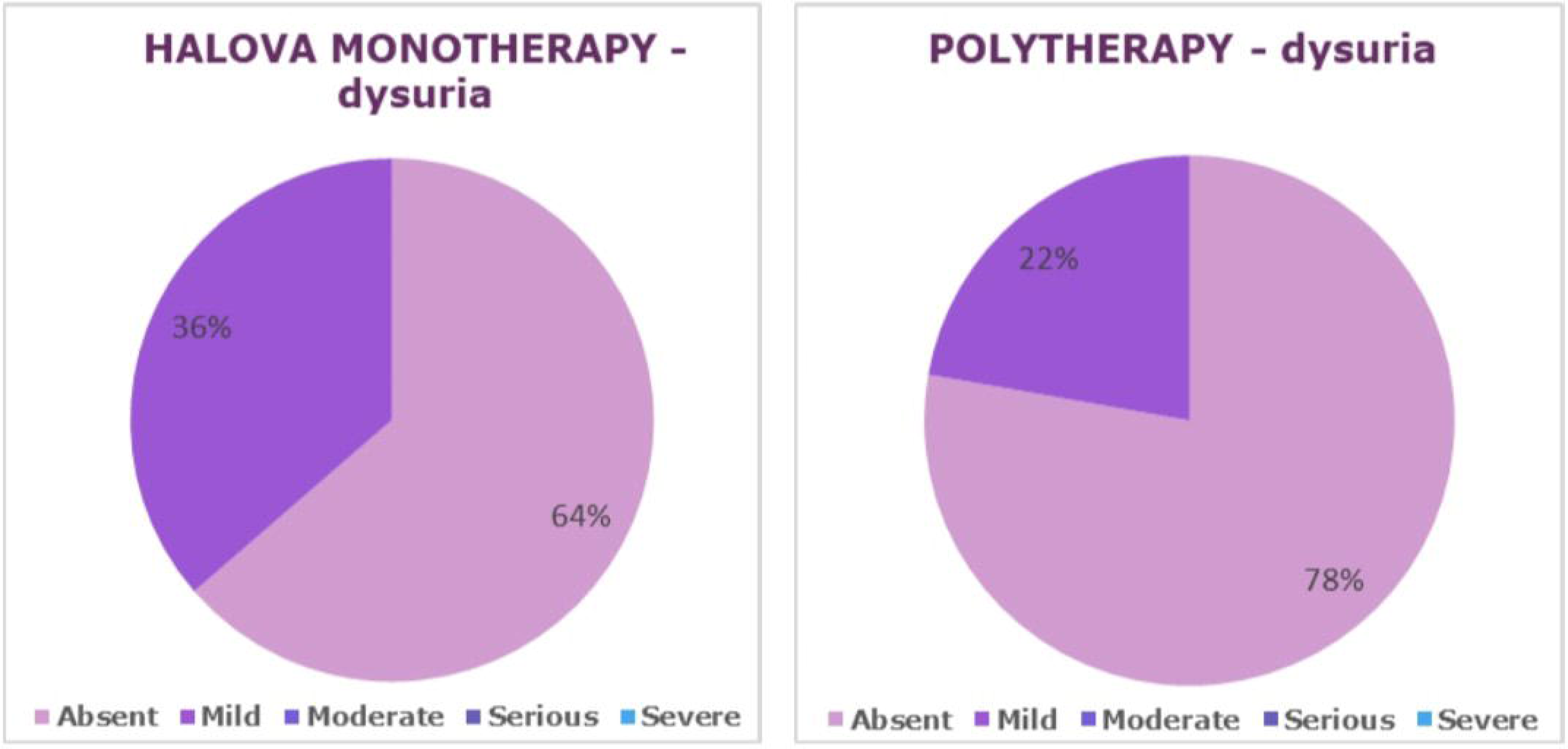

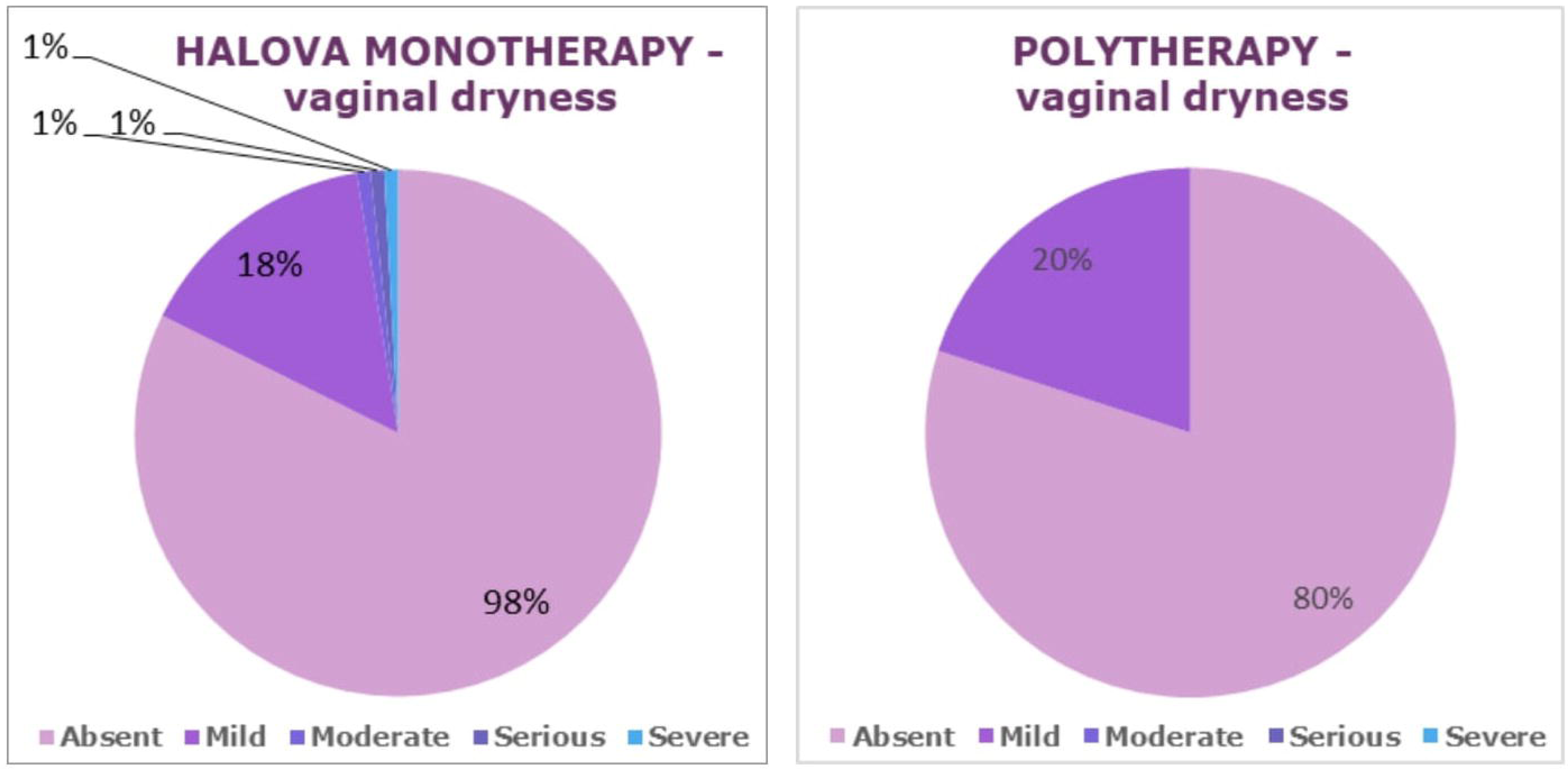

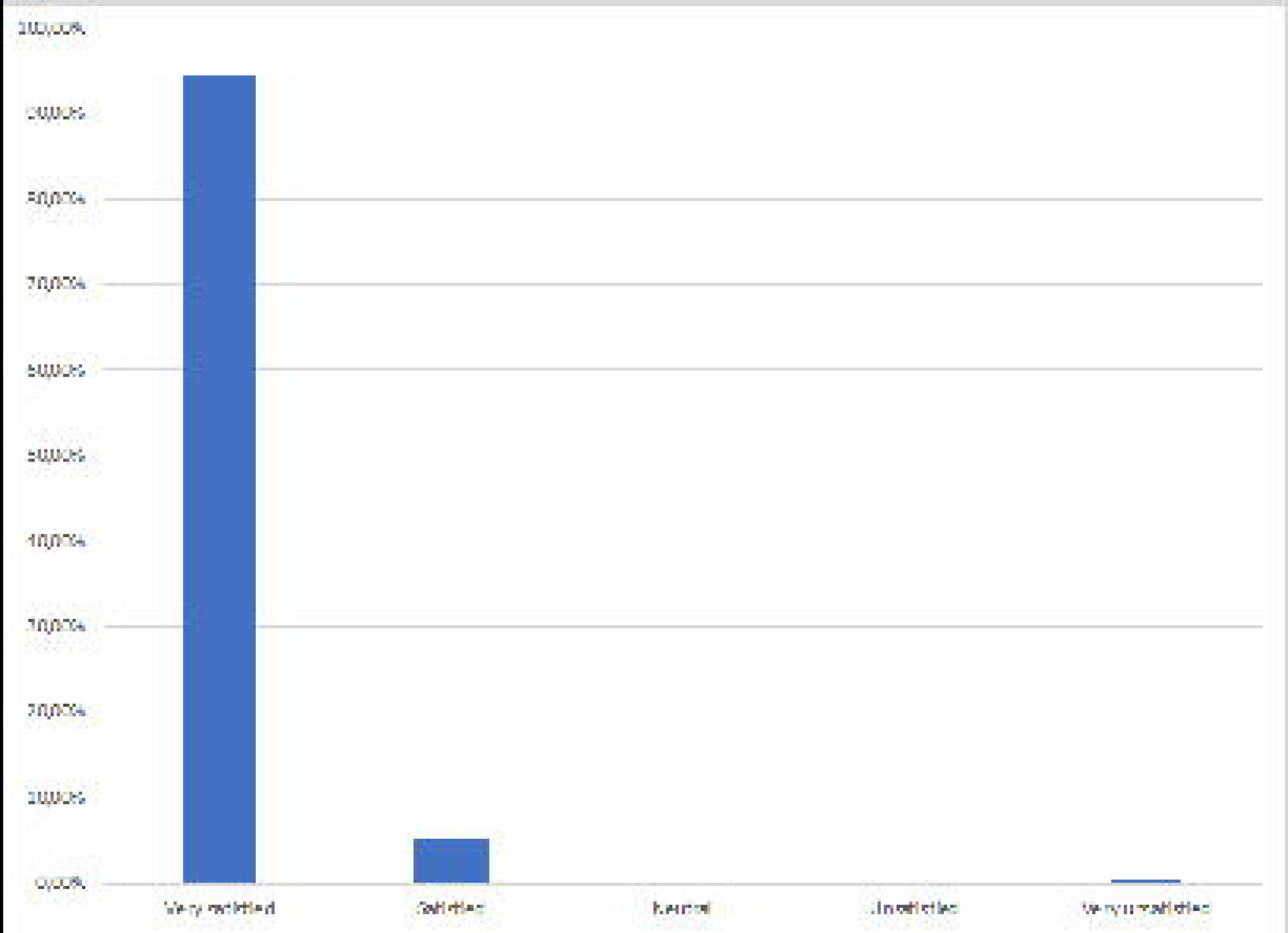

