## Supplementary figures and images for "Real-world performance and safety of Halova^®^ ovules in reducing the vaginal symptoms associated with vulvovaginal atrophy and postmenopausal sexual dysfunction" (from another version of this preprint: version 1, 2023-05-25)

### Supplemental Figure 5

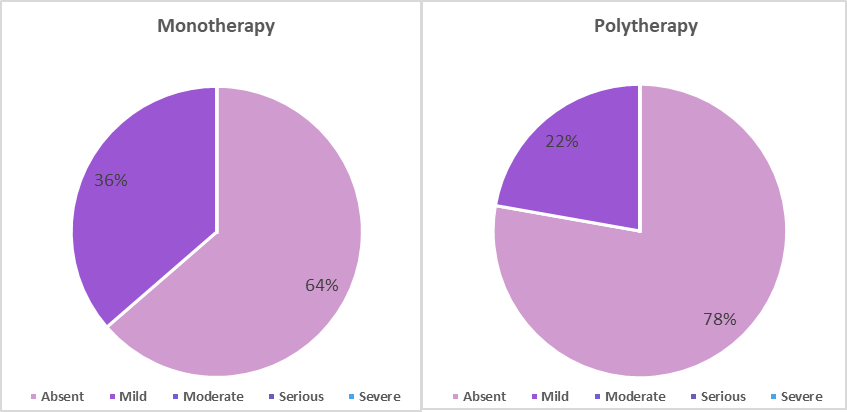

Figure 4: Dysuria symptoms – evaluation results at 30 days

### Supplemental Figure 6

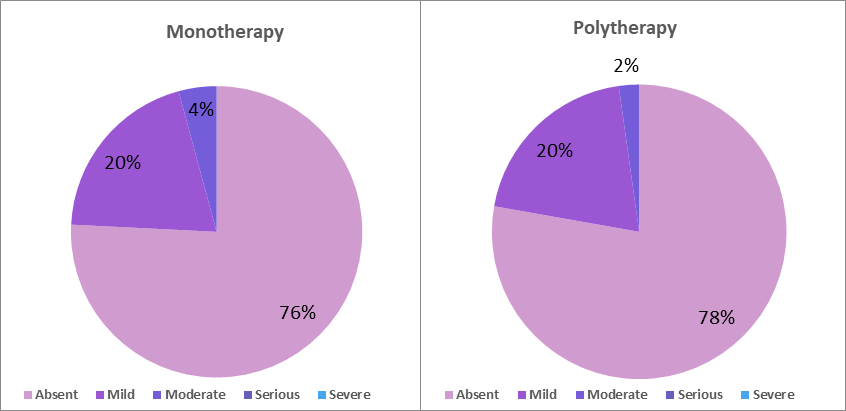

Figure 5: Endometrium thickness – evaluation results at 30 days

### Supplemental Figure 7

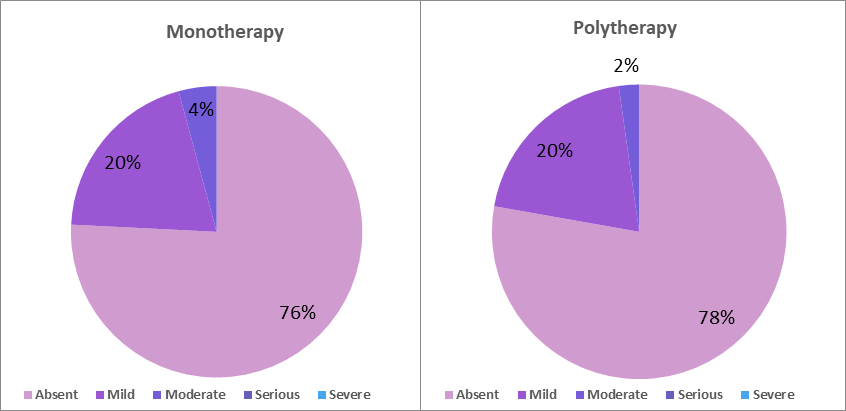

Figure 6: Vaginal dryness symptoms – evaluation results at 30 days
