## Supplemental Figure 8 for "Real-world performance and safety of Halova^®^ ovules in reducing the vaginal symptoms associated with vulvovaginal atrophy and postmenopausal sexual dysfunction" (from another version of this preprint: version 1, 2023-05-25)

Monotherapy

|  |  |
| --- | --- |
| Excellent | 94.38% |
| Highly effective | 5.06% |
| Effective | 0% |
| Minimally effective | 0% |
| Not effective | 0.56% |

Polytherapy

|  |  |
| --- | --- |
| Excellent | 85.71% |
| Highly effective | 14.29% |
| Effective | 0% |
| Minimally effective | 0% |
| Not effective | 0% |

Figure 7: Participant Satisfaction Score after using the medical device
